# Effects of transcranial direct current stimulation (tDCS) combined with cognitive therapy in individuals with cognitive impairment: a systematic review and meta-analysis

**DOI:** 10.64898/2026.04.26.26351755

**Authors:** Patricio Soto-Fernández, Lilian Toledo-Rodríguez, Alejandra Figueroa-Vargas, Paulo Figueroa-Taiba, Pablo Billeke

**Author notes:** Contact information: Patricio Soto-Fernández, Pablo Billeke.

## Abstract

**Background:** Cognitive impairment poses a significant challenge to healthcare systems worldwide, impacting patient autonomy, social participation, and quality of life, while placing a considerable burden on caregivers. Non-pharmacological interventions, particularly cognitive training and non-invasive brain stimulation, have emerged as promising therapeutic strategies.

**Objective:** This study aims to quantify the synergistic effects of transcranial direct current stimulation (tDCS) with cognitive training on cognitive function across a spectrum of pathologies that induce cognitive impairment.

**Methods:** We conducted a systematic review and meta-analysis following PRISMA guidelines. We searched PubMed for randomized controlled trials that investigated the effect of combined tDCS and cognitive training compared with cognitive training alone. The analysis was based on the GRADE framework for systematic reviews and meta-analyses.

**Results:** Across 27 studies including 1,012 participants, tDCS combined with cognitive training showed a small effect compared with cognitive training alone (SMD = 0.36, 95% CI: 0.15–0.56). The effect was found only immediately after the intervention and declined during follow-up.

**Conclusion:** tDCS combined with cognitive training may provide a small, short-term benefit for cognitive function, but high heterogeneity across studies and loss of effect at follow-up underscore the need for larger, better-standardized trials to clarify its clinical value.

**Highlights:**

- Combined tDCS and cognitive training produce a small but statistically significant short-term improvement in global cognitive performance.
- Effects attenuate over time, highlighting limited durability without sustained or maintenance interventions.
- High methodological heterogeneity and very low certainty of evidence limit broad clinical generalization.

## Introduction

Population aging is one of the most significant demographic transitions of the twenty-first century. Improvements in healthcare and living conditions have substantially increased life expectancy worldwide, leading to a rapidly growing proportion of older adults in the global population. This demographic shift has been accompanied by a rising prevalence of chronic age-related conditions, including both neurodegenerative and acquired disorders associated with cognitive impairment (1–4). Cognitive deficits are a hallmark of several of these conditions, including mild cognitive impairment (MCI), Alzheimer’s disease, Parkinson’s disease, stroke, and other neurological disorders, affecting core cognitive domains such as memory, attention, and executive functions that are critical for independent functioning (5,6).

The consequences of cognitive impairment extend beyond cognitive symptoms themselves. As cognitive decline progresses, individuals frequently experience reductions in functional independence, including difficulties performing activities of daily living and maintaining social engagement (7–9). These limitations often result in significant decreases in quality of life. In addition, caregivers commonly experience substantial emotional, physical, and financial burden associated with providing long-term support to individuals with cognitive impairment (10,11). At a societal level, the growing prevalence of cognitive disorders represents a major challenge for healthcare systems and social security structures, generating considerable economic costs related to medical care, long-term assistance, and caregiver support services (12).

From a neurobiological perspective, cognitive disorders share disruptions in cortical excitability and synaptic plasticity mechanisms. In particular, alterations in local neural activity and long-term potentiation (LTP)-like processes have been identified as key mechanisms underlying deficits in attention, memory, and executive function (13,14). Abnormalities in cortical excitability and impaired cortical plasticity have been consistently documented in individuals with mild cognitive impairment and Alzheimer’s disease, suggesting a common neurophysiological substrate that may represent a viable therapeutic target (15,16).

Despite extensive research, currently available pharmacological and behavioral interventions often produce limited or inconsistent improvements in cognitive functioning, and many fail to generate clinically meaningful benefits. These limitations have motivated the search for alternative therapeutic approaches that directly modulate neural activity and promote neuroplasticity (17,18). In this context, non-invasive brain stimulation techniques have emerged as promising tools for cognitive rehabilitation (19).

Transcranial direct current stimulation (tDCS) has gained increasing attention due to its capacity to modulate cortical excitability by applying weak electrical currents through scalp electrodes (20). tDCS offers several practical advantages for clinical use, including relatively low cost, easy administration, and minimal adverse effects (21). Importantly, this technique enables targeted modulation of cortical regions involved in specific cognitive processes, allowing for more localized neuromodulatory effects while potentially reducing systemic side effects commonly associated with pharmacological treatments, which may also present contraindications in certain patient populations (22–24). In parallel with advances in the understanding of the neurobiological mechanisms underlying cognition and brain plasticity, the number of clinical trials investigating tDCS as a therapeutic intervention for cognitive impairment has grown substantially over the past decade.

However, findings across studies remain heterogeneous and sometimes inconsistent (25). One important factor contributing to this variability is the diversity of stimulation protocols currently used, including differences in electrode montage, stimulation intensity, session duration, number of sessions, and concurrent cognitive interventions. This methodological heterogeneity complicates the identification of optimal stimulation parameters and limits the translation of research findings into standardized clinical protocols (26,27).

More recently, research has increasingly emphasized the importance of developing individualized neuromodulation approaches that account for participant-specific characteristics, including baseline cognitive performance, neurophysiological markers, and disease-specific mechanisms (28). A key element of this personalization is the timing and context of stimulation delivery, as the effects of neuromodulation depend on the state and activity of the targeted neural networks. In particular, for cognitive processes, stimulation may be more effective when administered concurrently with the relevant cognitive task. Within this framework, combining tDCS with cognitive training or cognitive therapy has been proposed as a promising strategy to enhance neuroplasticity by engaging task-relevant neural networks during stimulation (29–31).

Nevertheless, although an increasing number of clinical trials have investigated the combined use of tDCS and cognitive interventions in individuals with cognitive impairment, the magnitude and consistency of their effects remain unclear (32). Previous studies have reported mixed results, potentially due to differences in study design, stimulation parameters, patient populations, and outcome measures (33–35). A comprehensive synthesis of the current evidence is therefore needed to understand the effectiveness of these combined interventions better and to identify stimulation protocols that may yield the most robust cognitive benefits.

Therefore, the present systematic review and meta-analysis aim to evaluate the available evidence on the effects of tDCS combined with cognitive therapy on cognitive outcomes in individuals with cognitive impairment, and to identify stimulation parameters and intervention characteristics that may be associated with greater therapeutic efficacy.

## Methods

### Search and study selection

To determine the effect of adding tDCS to conventional therapy in individuals with cognitive impairment, we developed a systematic review and meta-analysis. The search strategy (Table S1) was structured around PICO parameters (36) and conducted using the MEDLINE database aligned with the Cochrane Library’s guidance on systematic review methodology (37). No date restrictions were applied in the search, allowing the inclusion of all relevant studies regardless of publication year. The results were screened and presented following the PRISMA diagram. Article selection and extraction were carried out by five independent researchers using the COVIDENCE platform. The inclusion and exclusion criteria are detailed in Table 1. Data extraction from the selected articles was then performed in an Excel spreadsheet for subsequent analysis.

**Table 1.** Inclusion and exclusion criteria based on PICO question.

| PICO parameters | Inclusion | Exclusion |
| --- | --- | --- |
| Population | Adults diagnosed with mild or moderate cognitive impairment of any etiology. | Studies involving participants with severe cognitive impairment or diagnosed dementia with advanced functional decline. |
| Intervention | Studies evaluating transcranial electrical stimulation (tES), specifically tDCS or tACS, combined with cognitive stimulation or cognitive training | Interventions using brain stimulation techniques other than tDCS or tACS (e.g., TMS, electroconvulsive therapy). |
| Comparator | Cognitive stimulation or cognitive training alone. | Studies not combining the electrical stimulation intervention with cognitive training or cognitive stimulation. |
| Outcome | Studies reporting at least one of the following:<br>- Clinical improvement<br>- Adverse events<br>- Improvement on functional scales<br>Improvement in independence in activities of daily living | Studies not reporting any of the predefined outcomes. |
| Type of studies | Randomized controlled trials or controlled clinical trials. | Absence of a comparison group receiving cognitive stimulation/training alone. Case reports, reviews, qualitative studies, conference abstracts, or protocols without available results. |

### Data Analysis

The extracted data were analyzed qualitatively and quantitatively. For the qualitative analysis, we focused on describing the characteristics of the populations included in the different studies, the stimulation protocols applied, and the specific brain regions targeted by the interventions. In contrast, the quantitative analysis examined outcomes related to improvements in cognitive performance, adverse events, and functional enhancements. These outcomes were synthesized through a summary of findings using GRADE methodology (38).

The quantitative analysis (meta-analysis) was performed using RevMan software, focusing on the different outcomes. For dichotomous outcomes, relative risk was calculated using the Mantel-Haenszel statistical method and a random-effects model. The results were then transformed into absolute values and summarized in the findings table. For continuous outcomes, the mean difference and standardized mean difference were calculated using the inverse-variance method within a random-effects model. The effect size was analyzed using the minimal important difference for the mean difference and Cohen’s coefficient for the standardized mean difference (SMD).

For each outcome, the effect size was calculated considering the six thresholds proposed in the gradebook (38). For outcomes expressed as relative effects, the thresholds were derived from health utility values. For outcomes measured with continuous scales and analyzed using mean differences, the thresholds were based on the minimal important difference. For outcomes analyzed using SMD, Cohen’s coefficient was used to establish the effect estimation thresholds.

### Risk of bias

The risk of bias was assessed using the Cochrane ROB-2 tool (39). Only studies selected for the quantitative analysis were evaluated. Two investigators independently reviewed each article, and in cases of disagreement, a third investigator was involved to resolve the differences.

### Quality of Evidence - GRADE

The certainty of the evidence was evaluated using the GRADE framework. The assessment considered the parameters of risk of bias, indirectness, inconsistency, and imprecision. This process followed the GRADE methodology (38), and the information for each outcome is summarized in the footnote of the Summary of Findings table.

## Results

The search was conducted in January 2025 and repeated in June 2025 to update the data. We identified 601 articles. After screening by title, abstract, and full text, a total of 68 articles were selected, of which 27 were selected for data extraction and quantitative analysis (see Figure 1). Among the selected studies for analysis, 8 studies were analyzed for MCI post stroke (30,31,40–45), 4 articles for Alzheimer’s dementia (46–49), 2 for cognitive impairment secondary to HIV (50,51), 1 for Multiple Sclerosis (52), 2 for primary progressive aphasia (PPA) (53,54), and 10 for MCI (55–64).

**Figure 1.**
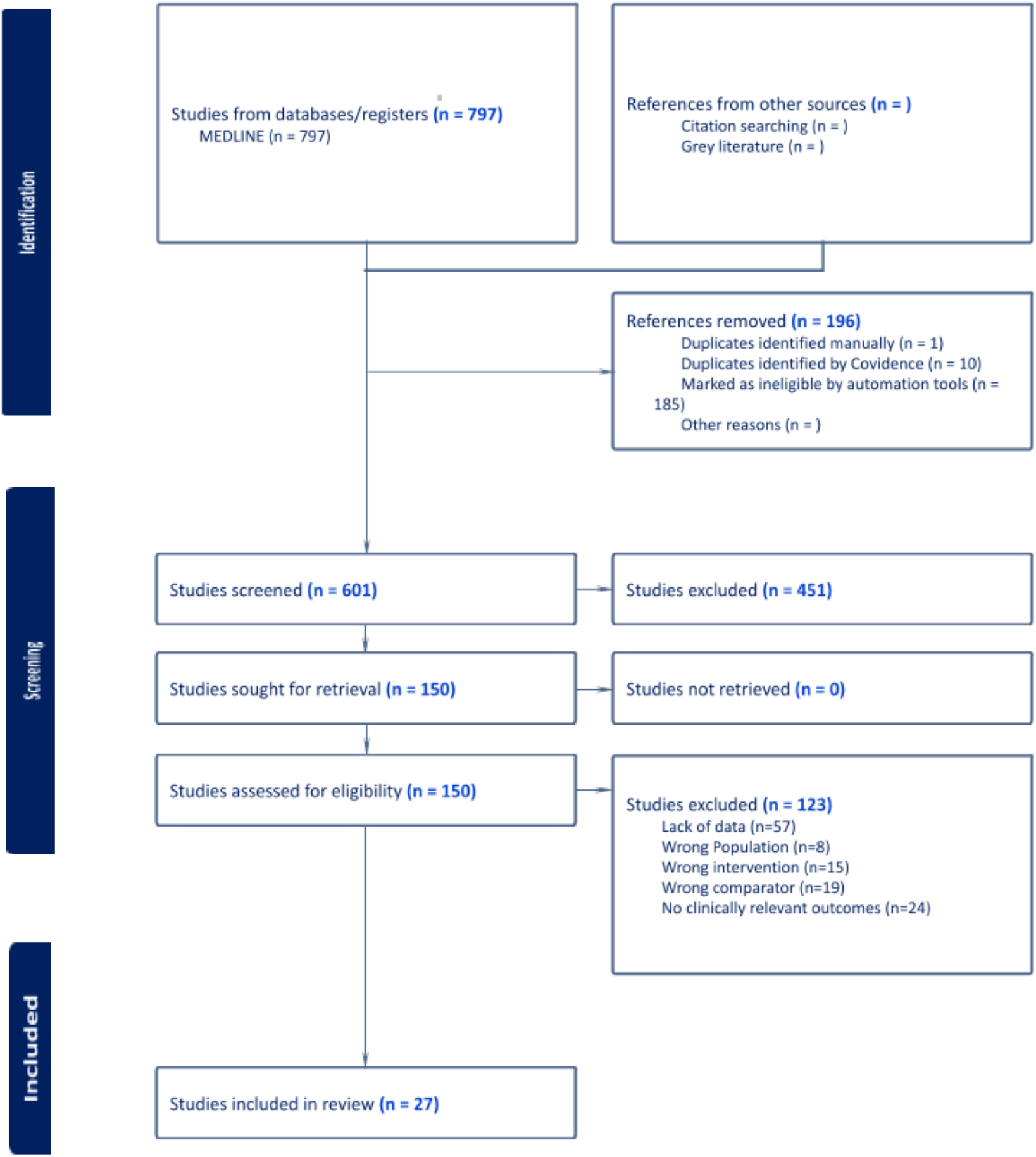
PRISMA flow diagram of literature search and study selection.

Risk of bias was assessed using the ROB-2 tool. Across the included studies, the most prominent potential bias arose from insufficient reporting of whether outcome adjudicators—specifically those conducting post-intervention assessments—were blinded to group allocation. In contrast, the remaining domains of bias were generally judged to be low risk. These patterns are summarized in Figure 2.

**Figure 2.**
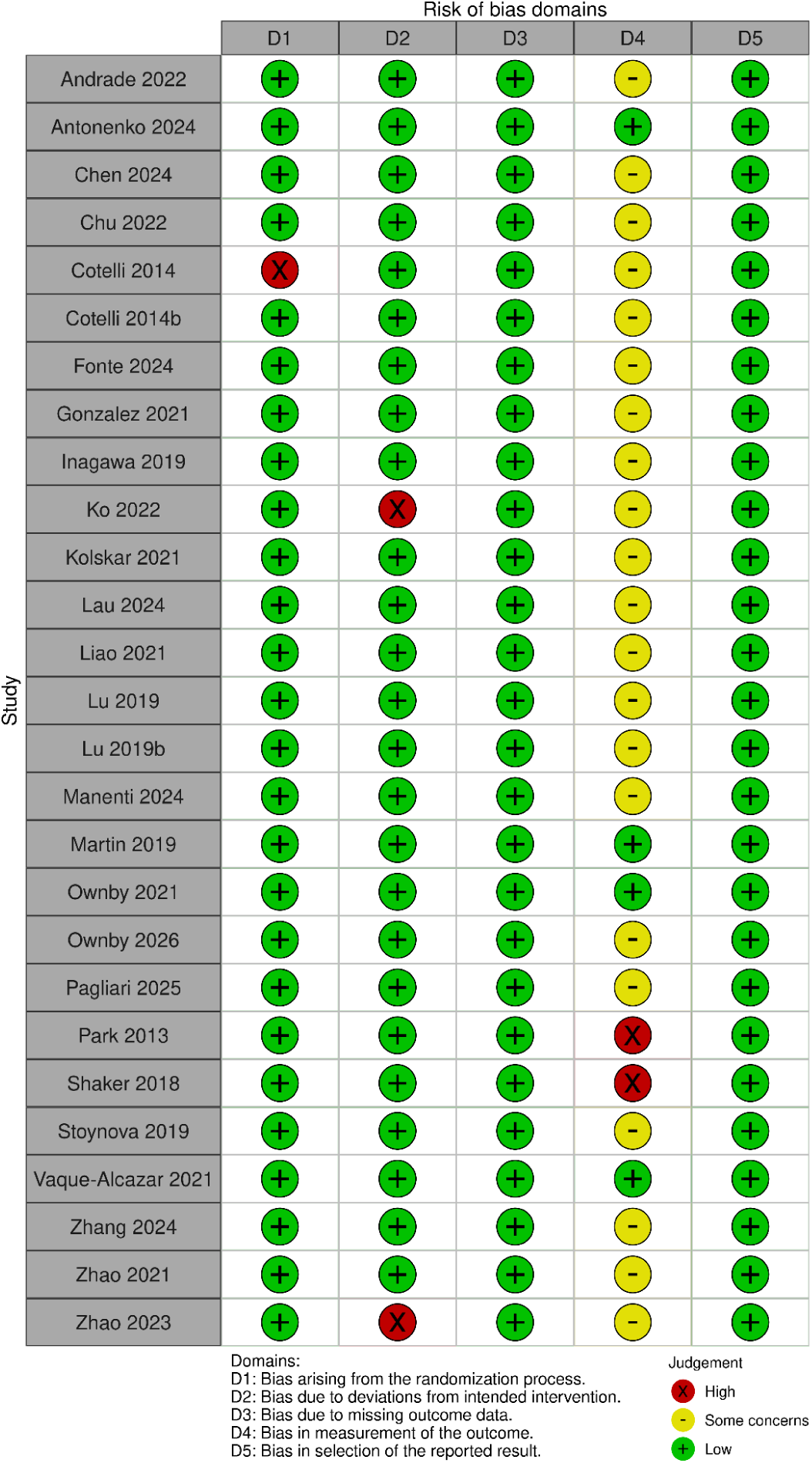
Risk of bias

Across the included studies, participant age ranged from 22 to 95 years; only the multiple sclerosis study selected individuals younger than 50, while all other samples consisted exclusively of adults over 50. Cognitive outcomes were assessed using instruments such as the Mini-Mental State Examination (MMSE), Montreal Cognitive Assessment (MoCA), Wechsler Adult Intelligence Scale (WAIS), Alzheimer’s Disease Assessment Scale – Cognitive Subscale (ADAS-Cog), California Verbal Learning Test (CVLT), and Paced Auditory Serial Addition Test (PASAT). For functional status or independence, the studies employed instruments such as the Activities of Daily Living (ADL) scale, the Instrumental Activities of Daily Living (IADL) scale, the Barthel Index, and the Functional Independence Measure (FIM), depending on the population and study design. Finally, not all articles reported adverse events; only 15 (30,31,40–45,50,52,55–58,64) studies reported data on this aspect. All this information is summarized in Table 2.

**Table 2.**
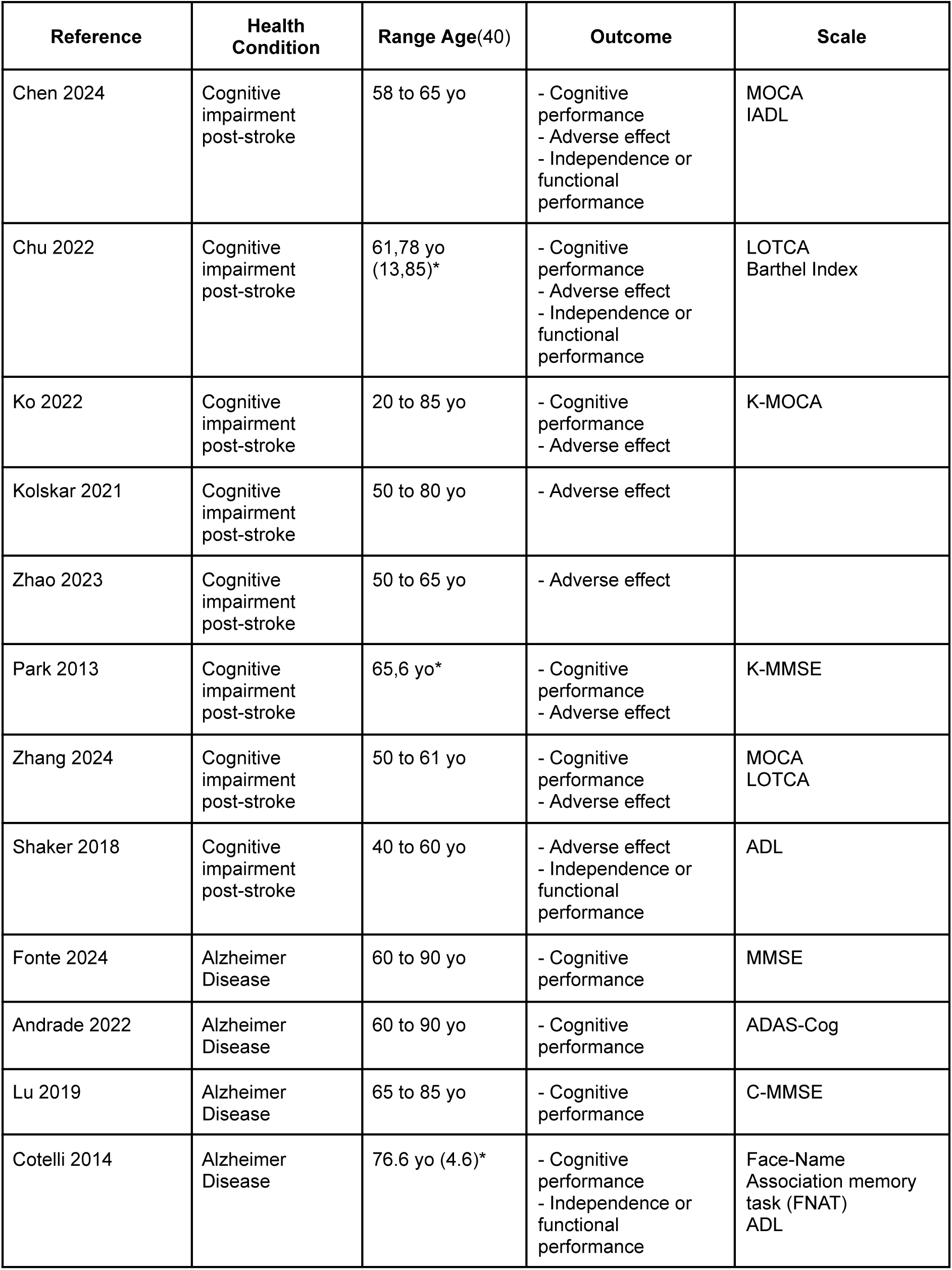
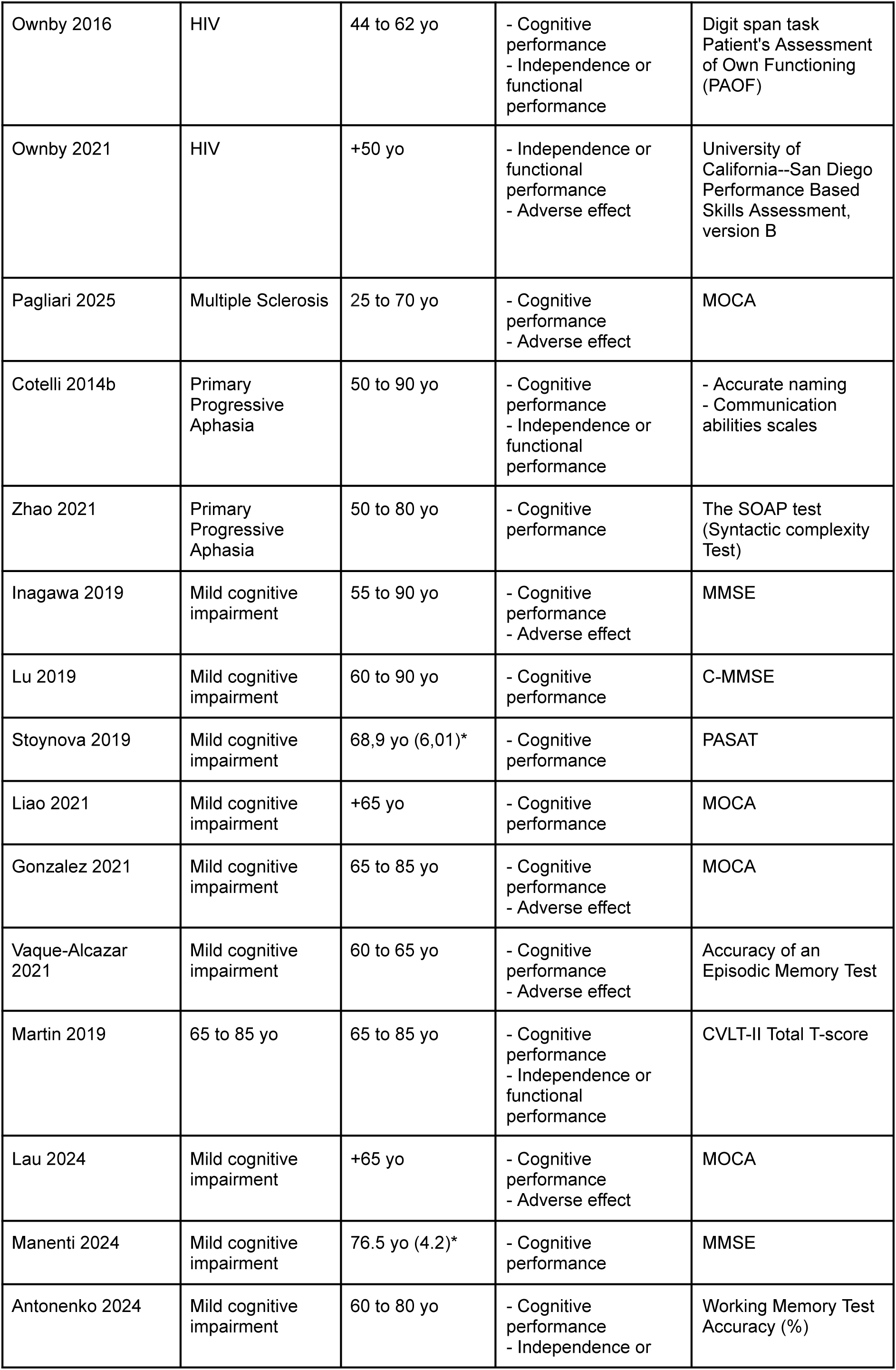
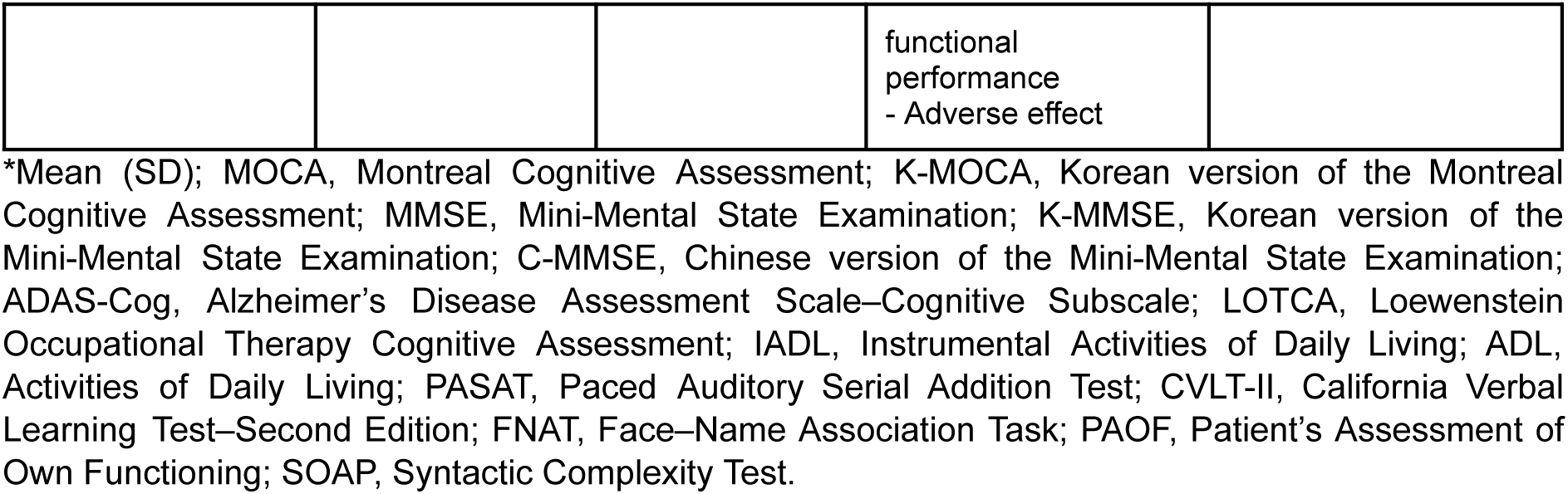
General characteristics of studies included in the analysis.

The stimulation parameters are described in Table 3. All included studies employed an anodal tDCS configuration, targeting the left dorsolateral prefrontal cortex (DLPFC), specifically positioned over F3 according to the International 10–20 EEG system. The stimulation intensity ranged from 1.0 to 2.0 mA, with 2.0 mA being the most frequently used parameter across protocols. Session duration ranged from 20 to 60 minutes (the most frequent was 20 minutes), and interventions were delivered at a median frequency of 3 sessions per week, over a median total intervention period of 4 weeks.

**Table 3.**
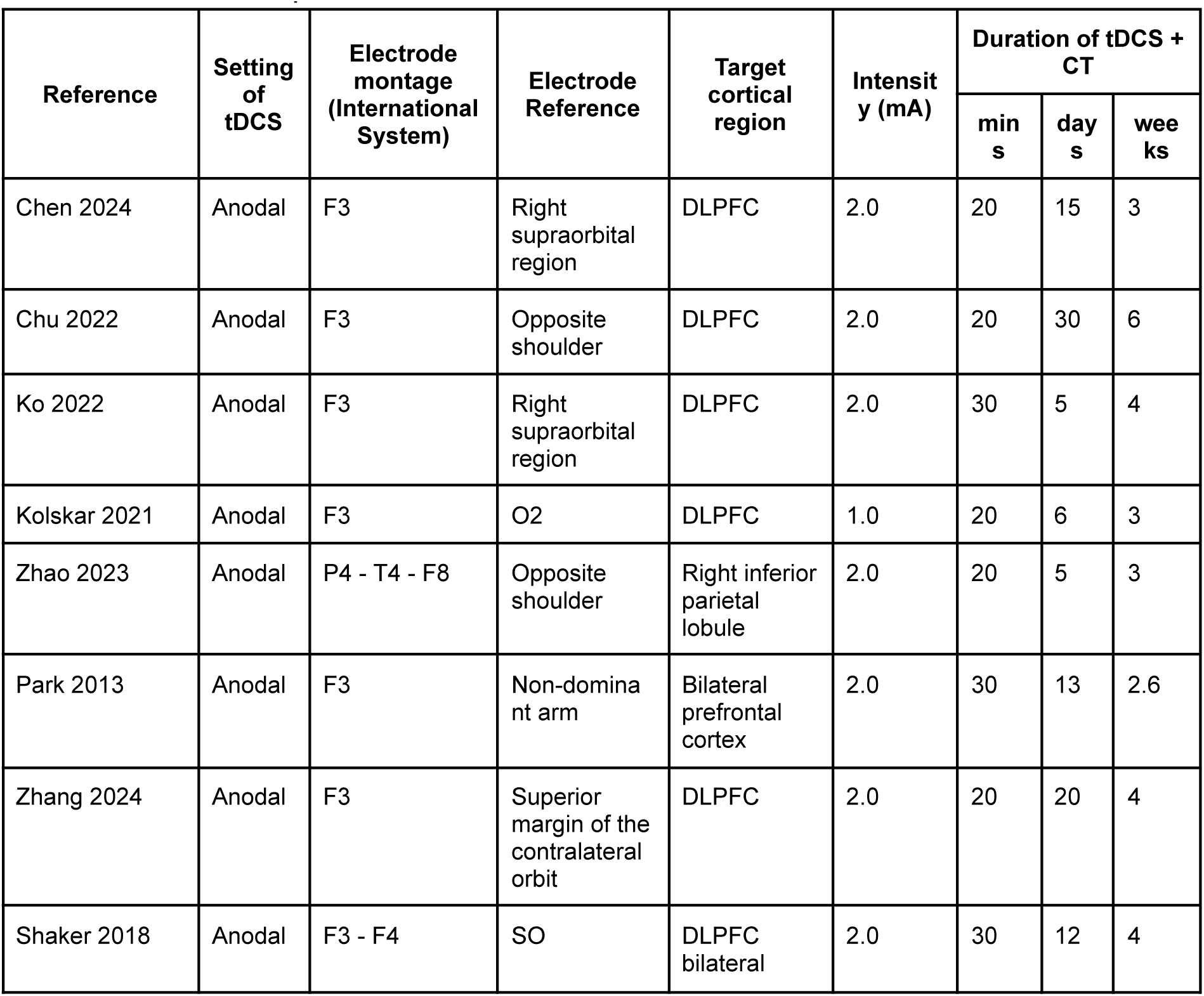
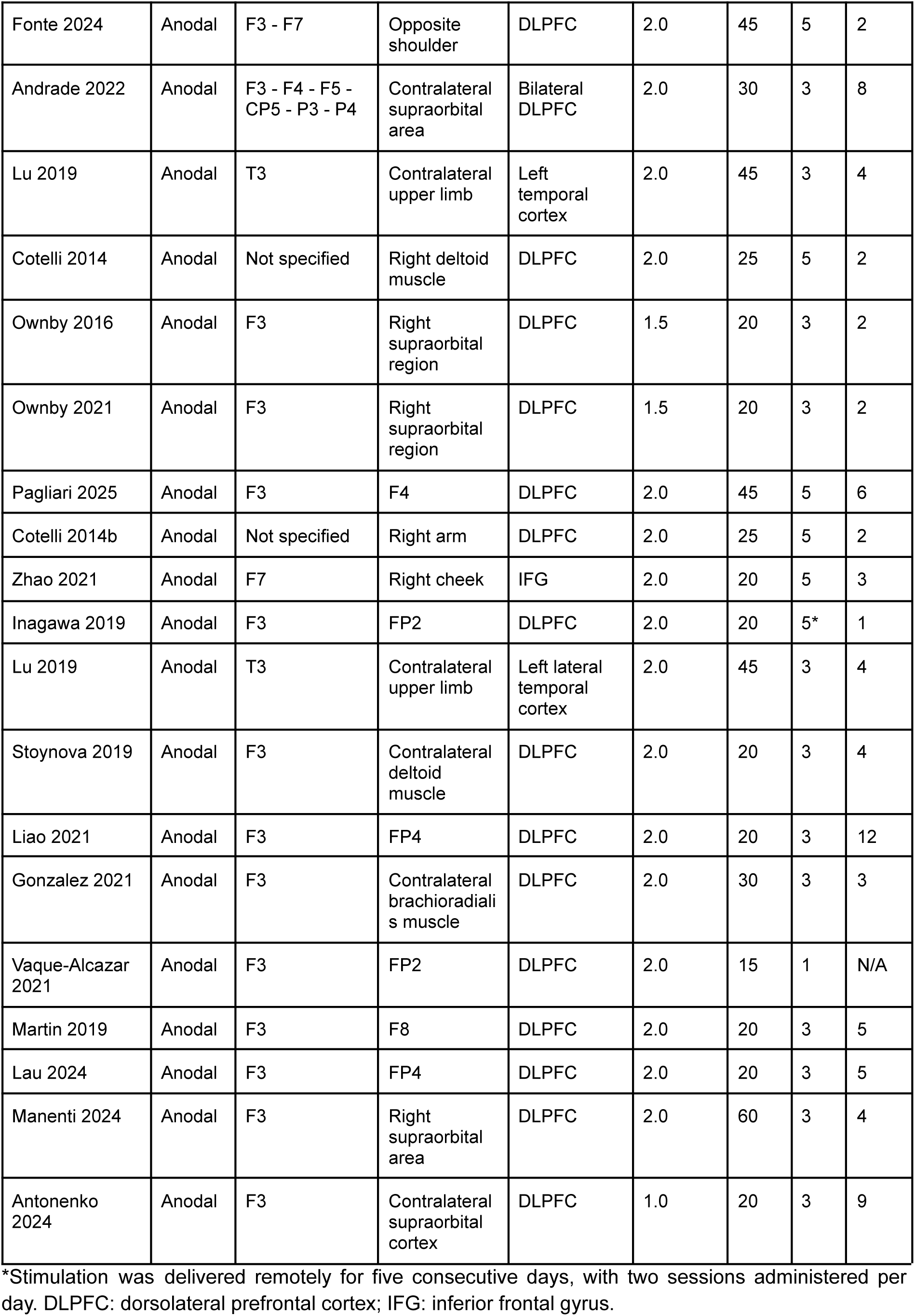
Stimulation parameters described in studies.

### Global Cognitive Performance

The long-term outcomes, stratified by the underlying etiology of cognitive deterioration, are presented in Table 4. Given the heterogeneity of measurement instruments across studies, the SMD was used to establish the effect size. Interpretation of the results followed Cohen’s conventional thresholds, considering an SMD of 0.2 or greater as indicative of a clinically meaningful effect.

**Table 4.**
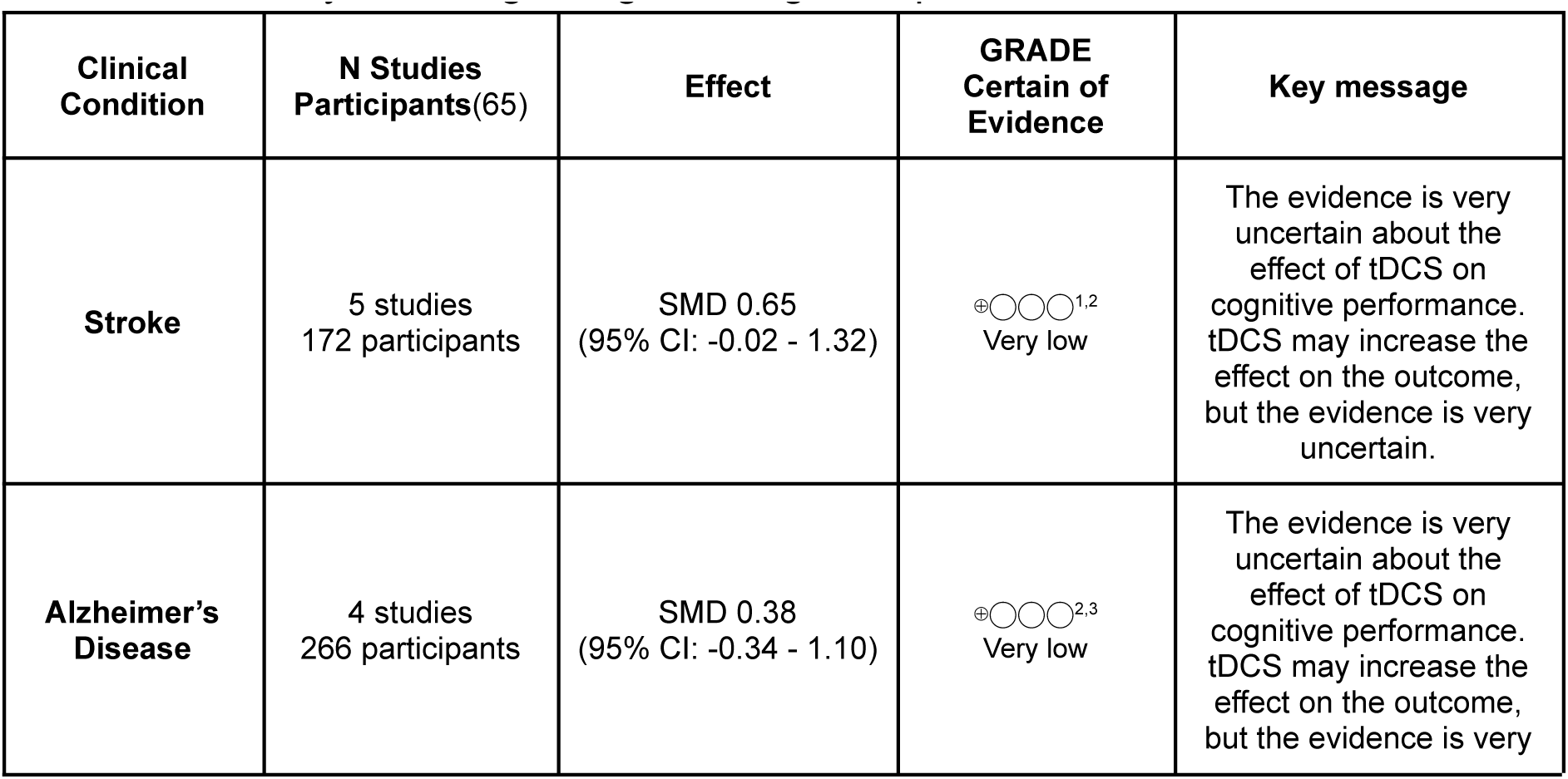
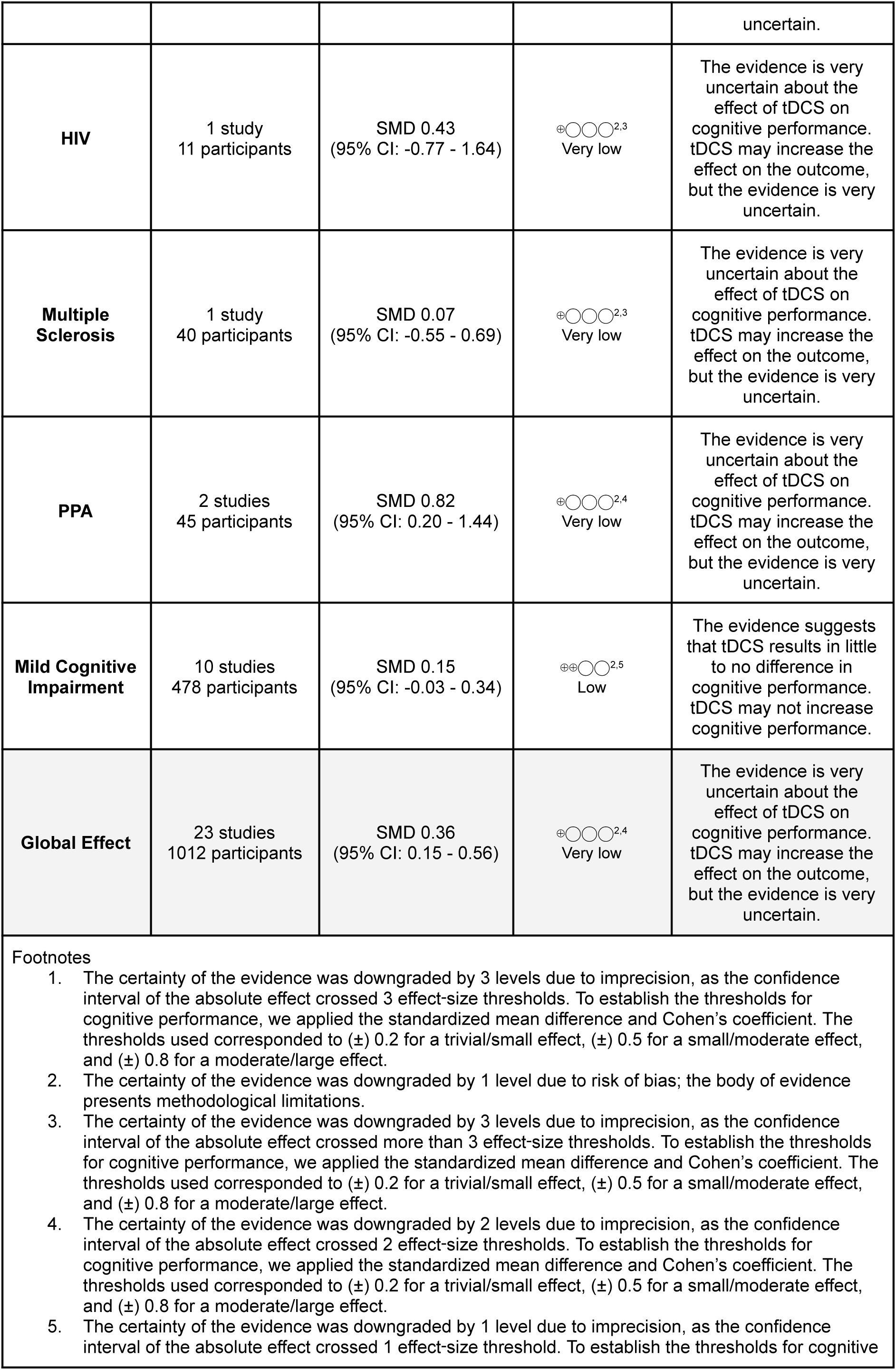
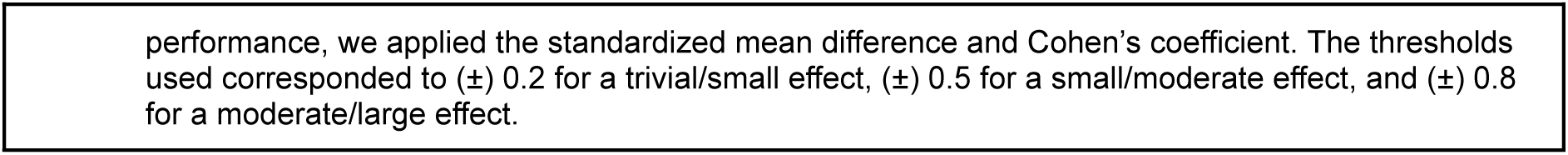
Summary of findings for global cognitive performance.

| Clinical Condition | N Studies<br>Participants(65) | Effect | GRADE<br>Certain of<br>Evidence | Key message |
| --- | --- | --- | --- | --- |
| **Stroke** | 5 studies<br>172 participants | SMD 0.65<br>(95% CI: -0.02 - 1.32) | ⊕○○○<sup>1,2</sup><br>Very low | The evidence is very uncertain about the effect of tDCS on cognitive performance. tDCS may increase the effect on the outcome, but the evidence is very uncertain. |
| **Alzheimer's Disease** | 4 studies<br>266 participants | SMD 0.38<br>(95% CI: -0.34 - 1.10) | ⊕○○○<sup>2,3</sup><br>Very low | The evidence is very uncertain about the effect of tDCS on cognitive performance. tDCS may increase the effect on the outcome, but the evidence is very uncertain. |
| **HIV** | 1 study<br>11 participants | SMD 0.43<br>(95% CI: -0.77 - 1.64) | ⊕○○○<sup>2,3</sup><br>Very low | The evidence is very uncertain about the effect of tDCS on cognitive performance. tDCS may increase the effect on the outcome, but the evidence is very uncertain. |
| **Multiple Sclerosis** | 1 study<br>40 participants | SMD 0.07<br>(95% CI: -0.55 - 0.69) | ⊕○○○<sup>2,3</sup><br>Very low | The evidence is very uncertain about the effect of tDCS on cognitive performance. tDCS may increase the effect on the outcome, but the evidence is very uncertain. |
| **PPA** | 2 studies<br>45 participants | SMD 0.82<br>(95% CI: 0.20 - 1.44) | ⊕○○○<sup>2,4</sup><br>Very low | The evidence is very uncertain about the effect of tDCS on cognitive performance. tDCS may increase the effect on the outcome, but the evidence is very uncertain. |
| **Mild Cognitive Impairment** | 10 studies<br>478 participants | SMD 0.15<br>(95% CI: -0.03 - 0.34) | ⊕⊕○○<sup>2,5</sup><br>Low | The evidence suggests that tDCS results in little to no difference in cognitive performance. tDCS may not increase cognitive performance. |
| **Global Effect** | 23 studies<br>1012 participants | SMD 0.36<br>(95% CI: 0.15 - 0.56) | ⊕○○○<sup>2,4</sup><br>Very low | The evidence is very uncertain about the effect of tDCS on cognitive performance. tDCS may increase the effect on the outcome, but the evidence is very uncertain. |

The overall pooled effect corresponded to a clinically meaningful improvement, with an SMD of 0.36 (95% CI: 0.15 to 0.56; Z=3.44; p=0.0006). The analysis showed I² = 55% (heterogeneity) and a subgroup comparison p-value of 0.28, indicating no statistical support for differences between subgroups. These subgroup-level distinctions can be further appreciated in the individual results stratified by the underlying cause of cognitive deterioration.

The largest effect was observed in the context of PPA rehabilitation, with an SMD of 0.82 (95% CI: 0.20 to 1.44; Z=2.61; p=0.009). This was followed by cognitive impairment secondary to stroke, showing an SMD of 0.65 (95% CI: −0.02 to 1.32; Z=1.91; p=0.06), and by HIV-associated cognitive impairment, with an SMD of 0.43 (95% CI: −0.77 to 1.64; Z=0.70; p=0.48). In contrast, mild cognitive impairment and multiple sclerosis did not demonstrate meaningful effects, with SMDs of 0.15 (95% CI: −0.03 to 0.34; Z=1.68; p=0.09) and 0.07 (95% CI: −0.55 to 0.69; Z=0.23; p=0.82), respectively.

Despite the favorable findings, the overall certainty of the evidence remains very low for the global outcome. This is primarily driven by the wide confidence intervals surrounding the point estimate, which indicate substantial imprecision and limit the robustness of the observed effect. The detailed GRADE certainty assessment for this outcome is presented in the Summary of Findings table (Table 4 and Figure S1).

In the domain of functional performance in activities of daily living, the overall pooled effect was clinically meaningful (SMD 0.41, CI95% -0.09 - 0.91; Z=1.61; p=0.11). However, substantial inconsistency is evident in the forest plot (see Figure S2). Studies conducted in Alzheimer’s disease, mild cognitive impairment, and primary progressive aphasia don’t demonstrate effects in the use of tDCS (SMD -0.25, -0.33, and 0.02). In contrast, a clear and favorable effect was observed exclusively in cases of post-stroke cognitive impairment (SMD 0.97 - CI95% 0.34 - 1.59). This pattern of results is likely attributable to heterogeneity (I^2^=73%) and is supported by statistical evidence of subgroup differences, suggesting that the functional benefits of tDCS may be condition-specific rather than consistent across etiologies.

Finally, the effect was examined across different follow-up periods, as illustrated in **Figure 3**. The results show that the magnitude of the global effect on cognitive performance progressively diminishes over time. In addition, the confidence intervals exhibit noticeable variation in their width, further reflecting the temporal instability of the estimated effect. This variability may be partly explained by the limited number of studies reporting follow-up beyond the post-intervention period, progressive attrition leading to smaller sample sizes over time (see Figure S3), the inherent heterogeneity of the underlying condition, or the instability of the intervention effect itself.

**Figure 3.**
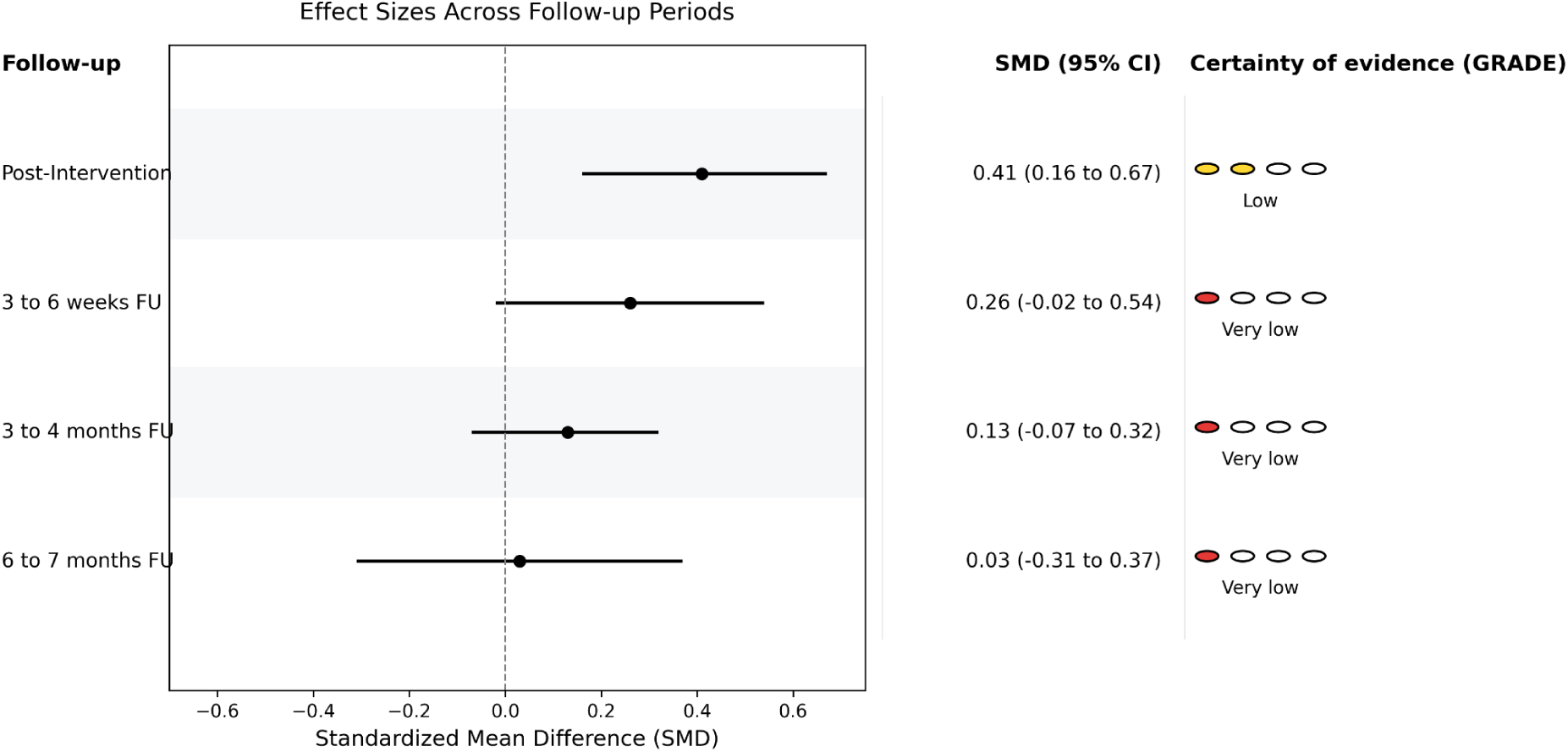
Estimated effect in different follow-ups. Effect sizes (standardized mean differences, SMD) across follow-up periods. Pooled estimates with 95% confidence intervals are shown for post-intervention (N = 999), 3 to 6 weeks follow-up (N = 365), 3 to 4 months follow-up (N = 433), and 6 to 7 months follow-up (N = 135). Certainty of evidence was assessed using the GRADE framework and is indicated for each time point.

In summary, combined anodal tDCS and cognitive therapy is associated with a small-to-moderate improvement in overall global cognitive performance. Functional improvements in activities of daily living appear condition-dependent, emerging primarily in post-stroke populations. However, the overall certainty of the evidence is very low, constrained by imprecision, heterogeneity, and risk-of-bias concerns, and the observed benefits attenuate over time.

## Discussion

The present systematic review and meta-analysis aimed to evaluate the effects of combining tDCS with cognitive training on cognitive outcomes in individuals with cognitive impairments across different etiologies. Our principal finding indicates that the combination yields a modest but statistically significant and likely clinically meaningful short-term improvement in global cognitive performance, although with substantial heterogeneity and low certainty of the evidence. Although this finding supports a potential adjunctive role for tDCS in cognitive rehabilitation, its interpretation should be tempered by the substantial heterogeneity observed across studies and the very low certainty of the evidence.

From a clinical perspective, these findings suggest that combined tDCS and cognitive therapy may be a useful adjunctive approach for cognitive rehabilitation, particularly in selected patient groups and when the stimulation is applied in close association with the cognitive task. The modest overall effect observed in our meta-analysis is consistent with previous work showing small but positive benefits of tDCS-based interventions on cognition, especially when stimulation parameters and behavioral training are well aligned (65,66). Importantly, prior studies have also suggested that the clinical impact of tDCS may depend on baseline cognitive resources, the target domain, and the specific neurological condition being treated, with some populations—such as stroke survivors—appearing more responsive than others (67,68). At the same time, the high heterogeneity across studies indicates that these benefits cannot yet be generalized to all patients or protocols, and that more standardized and etiologically stratified trials are needed to determine which individuals are most likely to benefit (69,70).

Several features of the included interventions may help explain the pattern of results. Considerable methodological heterogeneity (targets, intensities, session dosing, and cognitive task paradigms) and outcome heterogeneity (diverse neuropsychological instruments) contributed to between-study variability and limited the precision of pooled estimates (69). The subgroup signal favoring primary progressive aphasia and post-stroke conditions may reflect a better alignment between stimulation targets and the dysfunctional networks that are co-activated by language or domain-specific cognitive therapy. By contrast, mild cognitive impairment and multiple sclerosis trials—often characterized by broader cognitive phenotypes and variable disease mechanisms—did not show consistent benefits, which could also be due to smaller samples, different cognitive endpoints, or protocol mismatches.

The attenuation of effects over time raises questions about durability and the potential role of maintenance sessions or longer treatment courses to sustain gains. The rapid attenuation of effects observed at follow-up also highlights a critical limitation in current intervention designs: insufficient dosing and lack of maintenance protocols. The neuroplastic changes induced by tDCS and cognitive training may require sustained or repeated reinforcement to consolidate into long-term functional improvements (71,72). Future studies should systematically investigate dose-response relationships, including session frequency, total duration, and the potential role of booster sessions, to determine whether more durable cognitive gains can be achieved.

Given the very low certainty for the global cognitive outcome, routine adoption of combined tDCS and cognitive therapy cannot be broadly endorsed at this time. If considered, the application may be most justifiable in post-stroke cognitive impairment or primary progressive aphasia, ideally within research or quality-assurance frameworks, with careful target selection, standardized cognitive protocols, and systematic monitoring of outcomes and adverse events. Remote-supervised models may enhance access and adherence, but standardized safety reporting remains.

Personalization may be a key determinant of the clinical efficacy of non-invasive brain stimulation (28,73). In particular, targeting stimulation according to individual neurobiological profiles, including residual network activity and lesion-specific circuitry, may help optimize the effects of non-invasive brain stimulation. This is especially relevant in conditions such as post-stroke cognitive impairment, where preserved but functionally impaired networks may be more amenable to modulation. A key aspect relevant to tDCS personalization, and more broadly to non-invasive brain stimulation techniques, is the need for a precise alignment with specific clinical and cognitive therapeutic goals. Cognitive impairment does not present uniformly; rather, it manifests as distinct profiles across patients and pathologies (74–76). These profiles reflect individual patterns of brain network organization, function, and dynamics, underscoring the importance of tailoring interventions to each patient’s neurocognitive characteristics (77,78). Recent evidence suggests that specific cognitive processes, at the computational level, can be modulated, thereby improving the neural activity associated with them (79–81). For example, strong evidence across different pathologies has shown that specific signatures of electrical activity can be identified during working memory tasks (82–85). These features are crucial for enhancing the specificity of non-invasive brain stimulation during cognitive training, particularly when the intervention is tailored to the specific cognitive process (86). In this context, studies involving larger clinical samples, together with more precise physiological investigations, are urgently needed to strengthen the evidence base and to clarify how key brain functions and dynamics are associated with specific symptom profiles, thereby providing a stronger rationale for evaluating non-invasive brain stimulation therapies targeted to relevant clinical features.

A clear example of a neglected aspect in studies of cerebral functioning is the cerebellum. Emerging evidence suggests that cerebellar involvement should not be overlooked, given its role in cognitive and affective processing and its connections with frontoparietal control networks (71,72). In this context, biomarkers derived from functional MRI and EEG may provide valuable information to identify patients most likely to benefit from treatment by capturing baseline network integrity, task-related activation, and neurophysiological responsiveness. Such biomarker-guided approaches could move the field beyond a one-size-fits-all model toward more precise and mechanistically informed intervention strategies.

An additional critical factor that may account for variability in tDCS effects is state-dependency (28). The impact of stimulation is not only determined by anatomical targeting but also by the neural circuits’ ongoing activity at the time of stimulation. In this sense, the same stimulation protocol may produce different or even opposite effects depending on the baseline excitability and functional engagement of the targeted network (87). This principle reinforces the importance of delivering stimulation in conjunction with task-relevant cognitive engagement, as well as the potential value of monitoring neural states in real time to optimize intervention timing (88,89).

Importantly, translating these findings into real-world clinical settings requires careful consideration of feasibility, adherence, and scalability (90). While tDCS offers practical advantages, including low cost and ease of administration, implementing individualized, biomarker-informed protocols may pose logistical challenges that require advances in portable technologies and remote monitoring systems. Moreover, the transfer of cognitive improvements to functional gains in activities of daily living remains highly uncertain, due to the current lack of robust evidence supporting functional outcomes.

In conclusion, the present findings indicate that combining tDCS with cognitive training yields modest, short-lived improvements in cognitive performance, with effects highly dependent on clinical context, intervention design, and underlying neurobiological variability. Rather than reflecting a uniformly effective therapeutic approach, the current evidence suggests that the efficacy of combined neuromodulation strategies depends on alignment among stimulation parameters, targeted cognitive processes, and the integrity of the underlying neural networks. Moving forward, progress in this field will depend on integrating mechanistically informed, individualized intervention frameworks, supported by physiological biomarkers and computational models that capture the dynamic interplay between brain stimulation and cognitive function. Such approaches may enable a transition from heterogeneous, protocol-driven applications to precision neurorehabilitation strategies with greater clinical impact and durability.

## Data Availability

All data produced in the present work are contained in the manuscript

## Acknowledgements

This work was supported by Grants 1251073 (PB), 11261678 (AF-V) from the Fondo Nacional de Ciencia y Tecnología (FONDECYT) from Agencia Nacional de Investigación y Desarrollo (ANID). The funders had no role in study design, data collection, and analysis, the decision to publish, or the preparation of the manuscript.

---

**Table S1.**
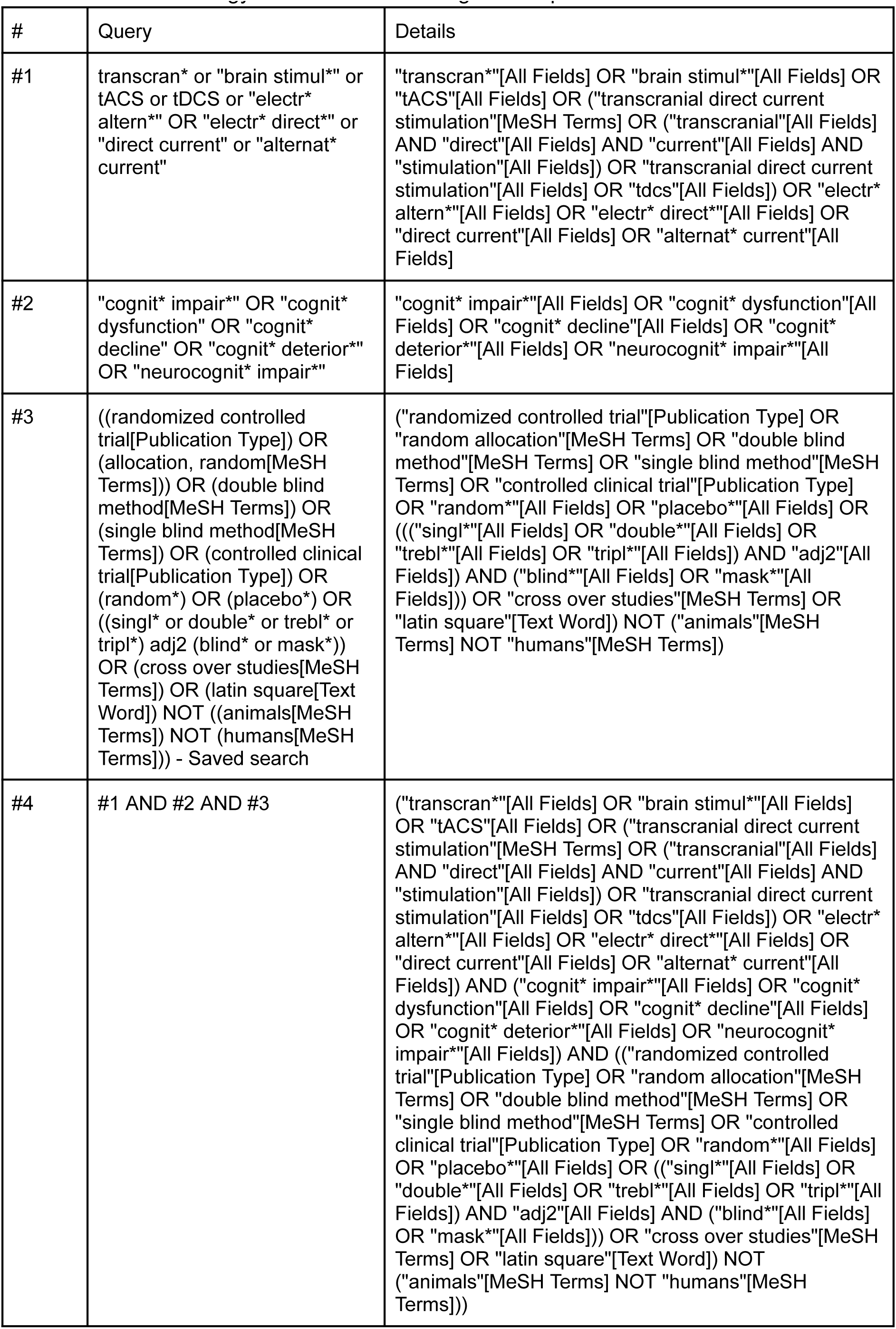
Search strategy for tDCS effect on cognitive impairment.

**Table S2.**
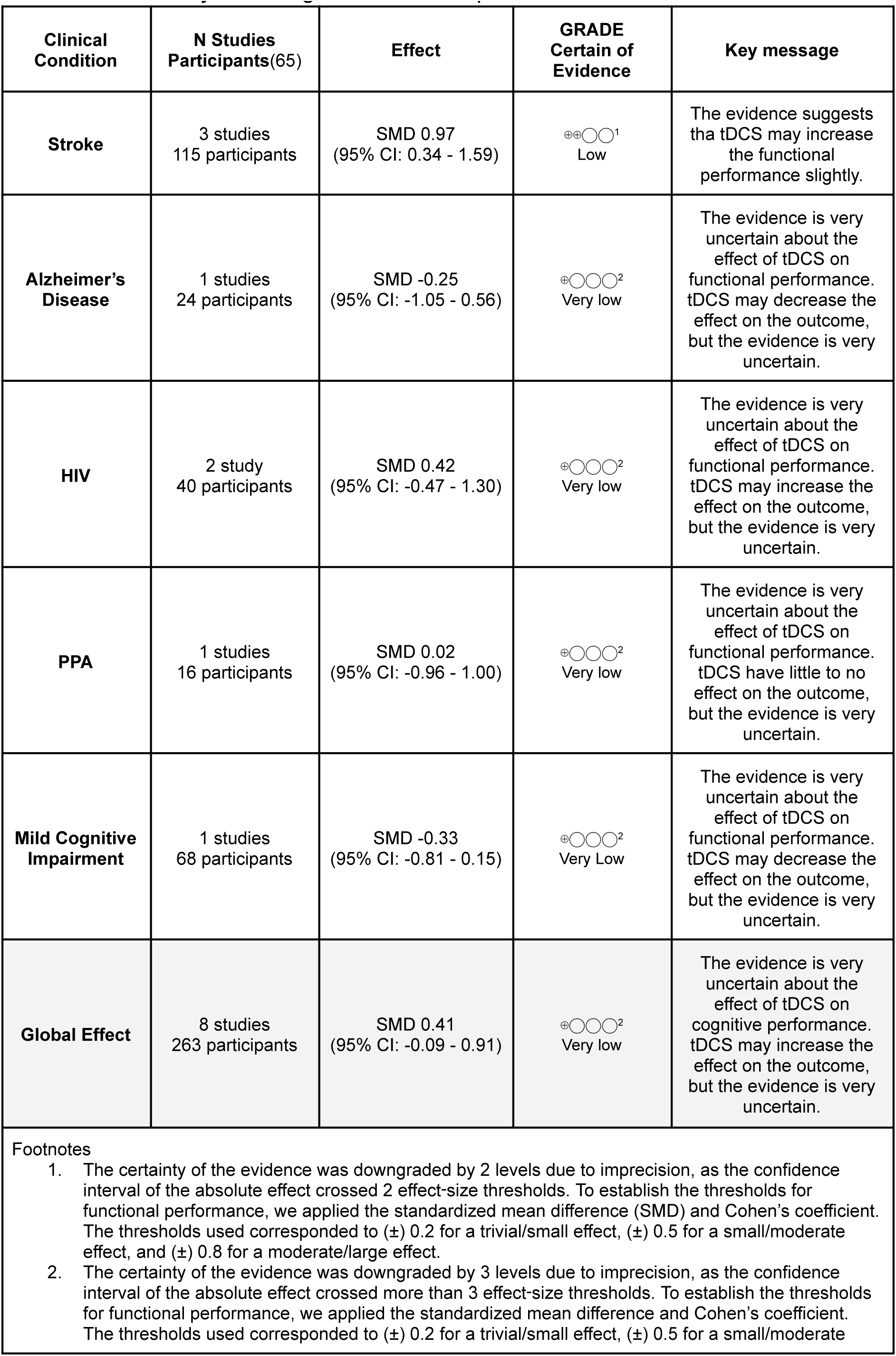
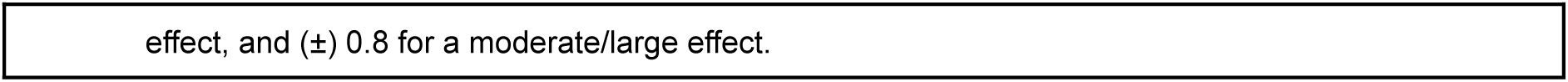
Summary of findings for functional performance.

| Clinical Condition | N Studies<br>Participants(65) | Effect | GRADE<br>Certain of<br>Evidence | Key message |
| --- | --- | --- | --- | --- |
| **Stroke** | 3 studies<br>115 participants | SMD 0.97<br>(95% CI: 0.34 - 1.59) | ⊕⊕○○<sup>1</sup><br>Low | The evidence suggests tha tDCS may increase the functional performance slightly. |
| **Alzheimer's Disease** | 1 studies<br>24 participants | SMD -0.25<br>(95% CI: -1.05 - 0.56) | ⊕○○○<sup>2</sup><br>Very low | The evidence is very uncertain about the effect of tDCS on functional performance. tDCS may decrease the effect on the outcome, but the evidence is very uncertain. |
| **HIV** | 2 study<br>40 participants | SMD 0.42<br>(95% CI: -0.47 - 1.30) | ⊕○○○<sup>2</sup><br>Very low | The evidence is very uncertain about the effect of tDCS on functional performance. tDCS may increase the effect on the outcome, but the evidence is very uncertain. |
| **PPA** | 1 studies<br>16 participants | SMD 0.02<br>(95% CI: -0.96 - 1.00) | ⊕○○○<sup>2</sup><br>Very low | The evidence is very uncertain about the effect of tDCS on functional performance. tDCS have little to no effect on the outcome, but the evidence is very uncertain. |
| **Mild Cognitive Impairment** | 1 studies<br>68 participants | SMD -0.33<br>(95% CI: -0.81 - 0.15) | ⊕○○○<sup>2</sup><br>Very Low | The evidence is very uncertain about the effect of tDCS on functional performance. tDCS may decrease the effect on the outcome, but the evidence is very uncertain. |
| **Global Effect** | 8 studies<br>263 participants | SMD 0.41<br>(95% CI: -0.09 - 0.91) | ⊕○○○<sup>2</sup><br>Very low | The evidence is very uncertain about the effect of tDCS on cognitive performance. tDCS may increase the effect on the outcome, but the evidence is very uncertain. |

1. The certainty of the evidence was downgraded by 2 levels due to imprecision, as the confidence interval of the absolute effect crossed 2 effect-size thresholds. To establish the thresholds for functional performance, we applied the standardized mean difference (SMD) and Cohen's coefficient. The thresholds used corresponded to (±) 0.2 for a trivial/small effect, (±) 0.5 for a small/moderate effect, and (±) 0.8 for a moderate/large effect. 2. The certainty of the evidence was downgraded by 3 levels due to imprecision, as the confidence interval of the absolute effect crossed more than 3 effect-size thresholds. To establish the thresholds for functional performance, we applied the standardized mean difference and Cohen's coefficient. The thresholds used corresponded to (±) 0.2 for a trivial/small effect, (±) 0.5 for a small/moderate
effect, and $(\pm) 0.8$ for a moderate/large effect.

**Figure S1.**
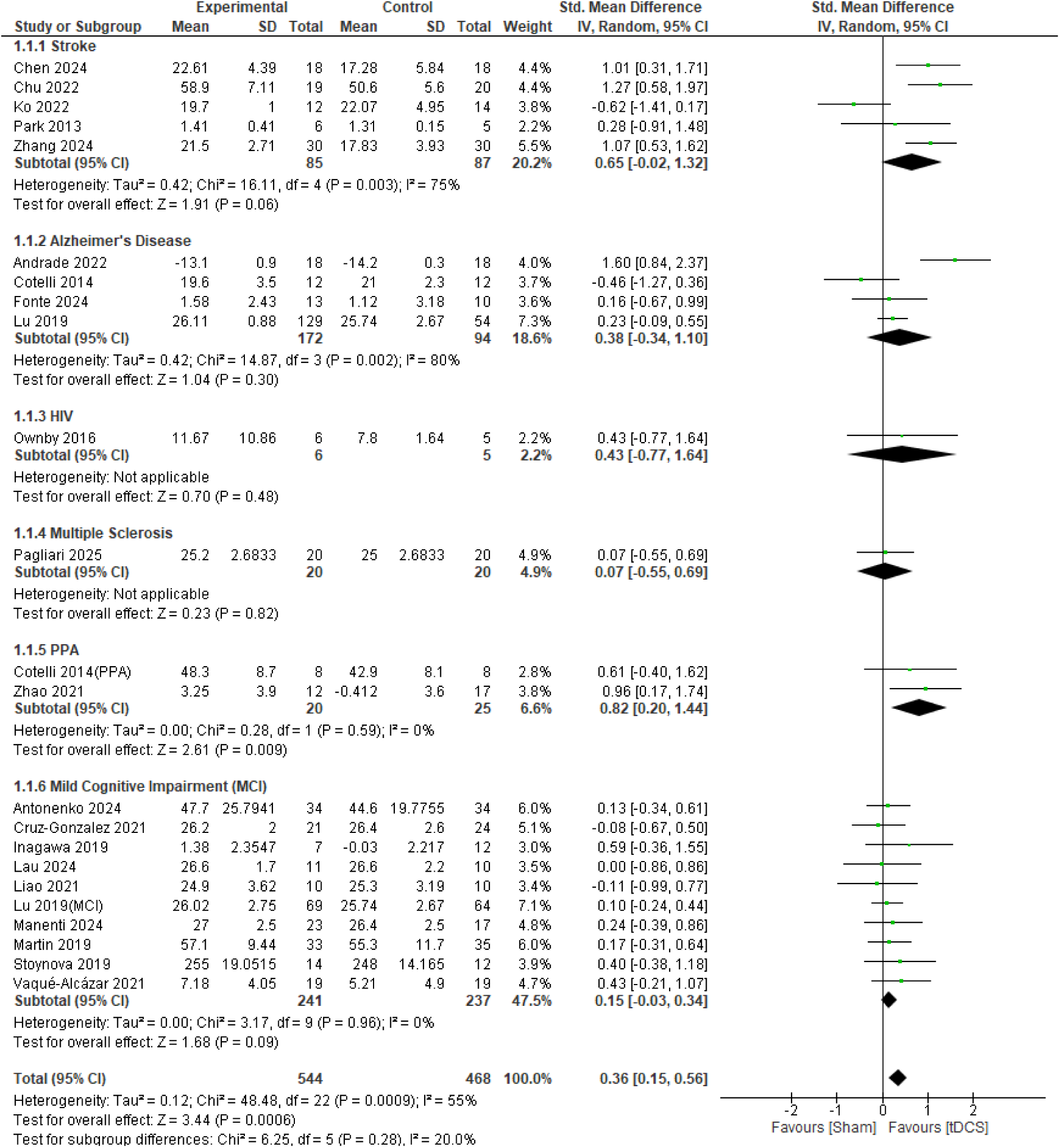
Forest plot for cognitive performance.

**Figure S2.**
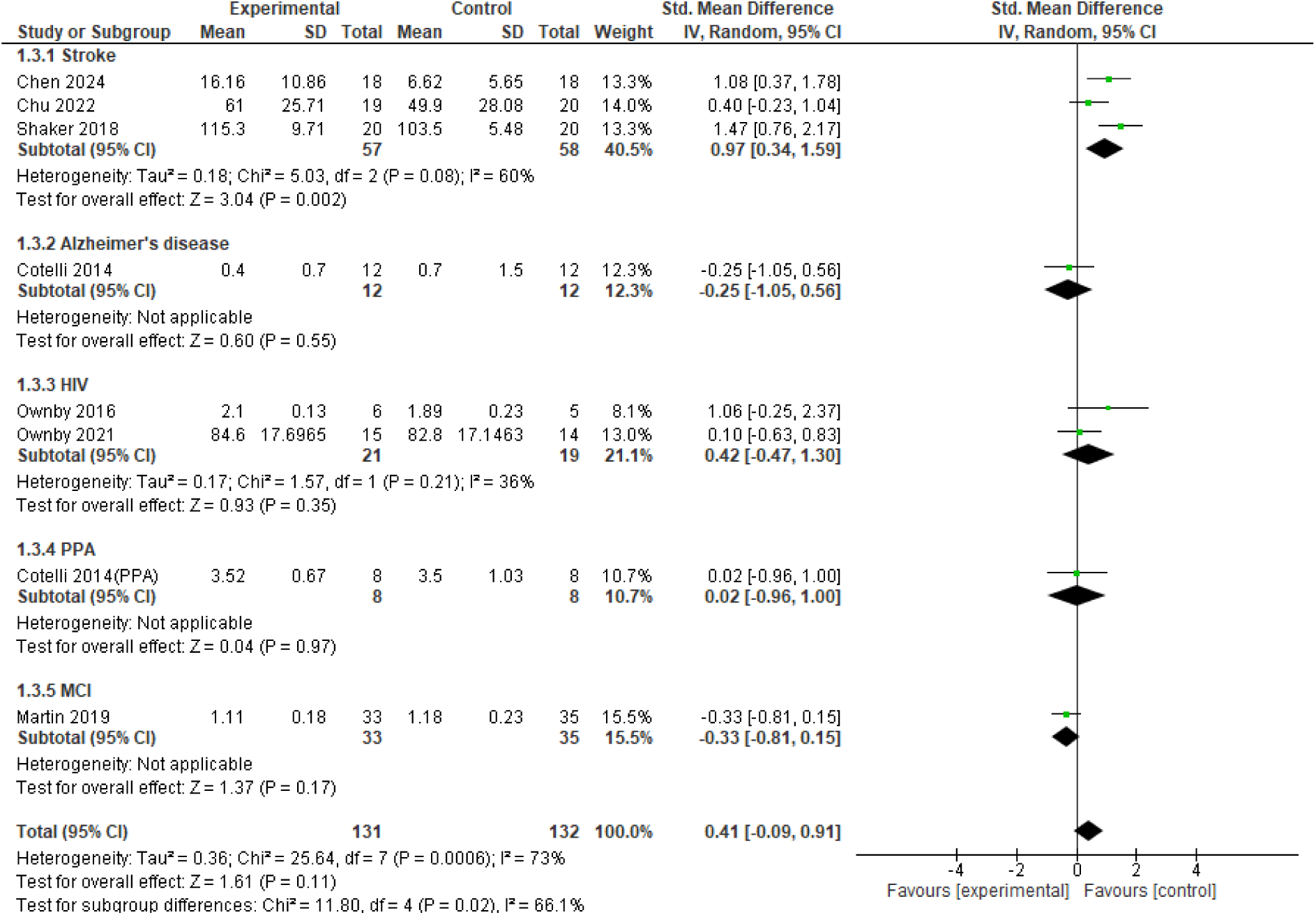
Forest plot for functional performance in activities of daily living.

**Figure S3.**
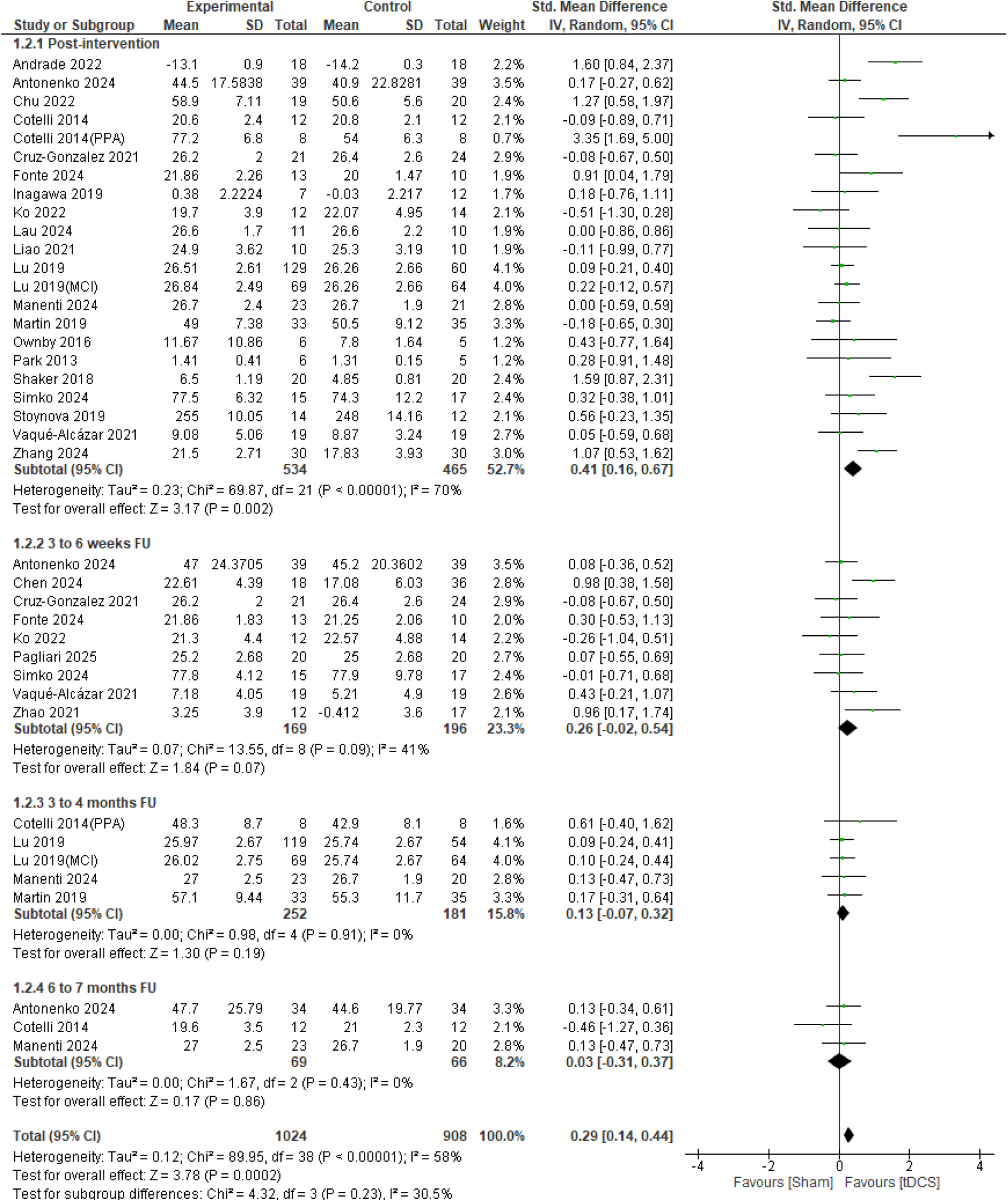
Effect of tDCS in cognitive performance in different follow-up.

